# Modeling the progression of SARS-CoV-2 infection in patients with COVID-19 risk factors through predictive analysis

**DOI:** 10.1101/2020.07.14.20154021

**Authors:** Juan Alonso Leon-Abarca, Esdras Ismael Musayón

## Abstract

With almost a third of adults being obese, another third hypertense and almost a tenth affected by diabetes, Latin American countries could see an elevated number of severe COVID-19 outcomes. We used the Open Dataset of Mexican patients with COVID-19 suspicion who had a definite RT-PCR result to develop a statistical model that evaluated the progression of SARS-CoV-2 infection in the population. We included patients of all ages with every risk factor provided by the dataset: asthma, chronic obstructive pulmonary disease, smoking, diabetes, obesity, hypertension, immunodeficiencies, chronic kidney disease, cardiovascular diseases, and pregnancy. The dataset also included an unspecified category for other risk factors that were not specified as a single variable. To avoid excluding potential patients at risk, that category was included in our analysis. Due to the nature of the dataset, the calculation of a standardized comorbidity index was not possible. Therefore, we treated risk factors as a categorical variable with two categories: absence of risk factors and the presence of at least one risk factor in accordance with previous epidemiological reports. Multiple logistic regressions were carried out to associate sex, risk factors, and age as a continuous variable (and the interaction that accounted for increasing diseases with older ages); and SARS-CoV-2 infection as the dependent zero-one binomial variable. Post estimation predictive marginal analysis was performed to generate probability trends along 95% confidence bands. This analysis was repeated several times through the course of the pandemic since the first record provided in their repository (April 12, 2020) to one month after the end of the state of sanitary emergency (the last date analyzed: June 27, 2020). After processing, the last measurement included 464,389 patients.

The baseline analysis on April 12 revealed that people 35 years and older with at least one risk factor had a lower risk of SARS-CoV-2 infection in comparison to patients without risk factors (Figure 1). One month before the end of the nationwide state of emergency this age threshold was found at 50 years (May 2, 2020) and it shifted to 65 years on May 30. Two weeks after the end of the public emergency (June 13, 2020) the trends converged at 80 years and one week later (June 27, 2020) every male and female patient with at least one risk factor had a higher risk of SARS-CoV-2 infection compared to people without risk factors. Through the course of the COVID-19 pandemic, all four probability curves shifted upwards as a result of progressive disease spread.

In conclusion, we found our model could monitor accurately the probability of SARS-CoV-2 infection in relation to age, sex, and the presence of at least one risk factor. Also, because the model can be applied to any particular political region within Mexico, it could help evaluate the contagion spread in specific vulnerable populations. Further studies are needed to determine the underlying nature of the mechanisms behind such observations.

---

With almost a third of adults being obese **(1)**, another third hypertense **(2)** and almost a tenth affected by diabetes **(3)**, Latin American countries could see an elevated number of severe COVID-19 outcomes. However, the estimation of precise SARS-CoV-2 figures and population-based analysis with individual data has been delayed by national policies which restricted access to anonymized patient data, with only Mexico, Peru and Colombia presenting large datasets in their respective national digital repositories.

In Mexico, primary healthcare centers take a nasopharyngeal swab for SARS-CoV-2 RT-PCR to every patient presenting respiratory distress or severe acute respiratory infections and one of every ten non severe ambulatory suspected cases who had symptoms related to COVID-19 in the last seven days in order to obtain representative figures of the pandemic in their country **(4)**. Because of that, we used the Open Dataset of Mexican patients with COVID-19 suspicion who had a definite RT-PCR result to develop a statistical model that evaluated the progression of SARS-CoV-2 infection in the population. We included patients of all ages with every risk factor provided by the dataset: asthma, chronic obstructive pulmonary disease, smoking, diabetes, obesity, hypertension, immunodeficiencies, chronic kidney disease, cardiovascular diseases and pregnancy. The dataset also included an unspecified category for other risk factors that were not specified as a single variable. To avoid excluding potential patients at risk, that category was included in our analysis. Due to the nature of the dataset the calculation of a standardized comorbidity index was not possible. Therefore, we treated risk factors as a categorical variable with two categories: "absence of risk factors" and "presence of at least one risk factor" in accordance to previous epidemiological reports **(5)**. Multiple logistic regressions were carried out to associate sex, risk factors and age as a continuous variable (and the interaction that accounted for increasing diseases with older ages); and SARS-CoV-2 infection as the dependent zero-one binomial variable. Post estimation predictive marginal analysis was performed to generate probability trends along 95% confidence bands. This analysis was repeated several times through the course of the pandemic since the first record provided in their repository (April 12, 2020) to one month after the end of the state of sanitary emergency (last date analyzed: June 27, 2020). After processing, the last measurement included 464,389 patients.

The characteristics of the study population showed that the mean age was 42.6 years old (Standard deviation = 16.6 years) and the male-to-female ratio was 1.025. Half of the patients had no risk factors (56.92%) while a quarter had only one (25.8%). The most common risk factors found in adults (18 years old and older) were hypertension (17.08%), obesity (17.08%) and diabetes (13.11%); and the most common risk factors present in people younger than 18 years were immunodeficiencies (3.86%), asthma (3.83%) and obesity (3.14%).

The baseline analysis at April 12 revealed that people 35 years and older with at least one risk factor had a lower risk of SARS-CoV-2 infection in comparison to patients without risk factors **(Figure 1)**. One month before the end of the nationwide state of emergency this age threshold was found at 50 years (May 2, 2020) and it shifted to 65 years at May 30. Two weeks after the end of the public emergency (June 13, 2020) the trends converged at 80 years and one week later (June 27, 2020) every male and female patient with at least one risk factor had a higher risk of SARS-CoV-2 infection compared to people without risk factors. Through the course of the COVID-19 pandemic, all four probability curves shifted upwards as a result of progressive disease spread. These findings might be explained by the increased awareness of vulnerability of both people with risk factors and the elderly population who might be more likely to comply with the official "stay at home" recommendations along with family members or caregivers that could have contributed by keeping them safer in enclosed settings. However, some official policies could have unintentionally accelerated the spread of SARS-CoV-2 infection in the older population, such as the recommendation of increased interaction with the elderly to avoid mental health issues as a result of the voluntary confinement in May 26 **(6)**. The Mexican National Institute of Geriatry has issued a continuously updated and extensive guideline intended to the general public on how to prevent SARS-CoV-2 contagion to elder people **(7)**. However, additional measures to prevent infection might be needed, because as the pandemic spreads, older demographic groups could take the hardest toll of COVID-19 complications.

**Figure Image 1.**
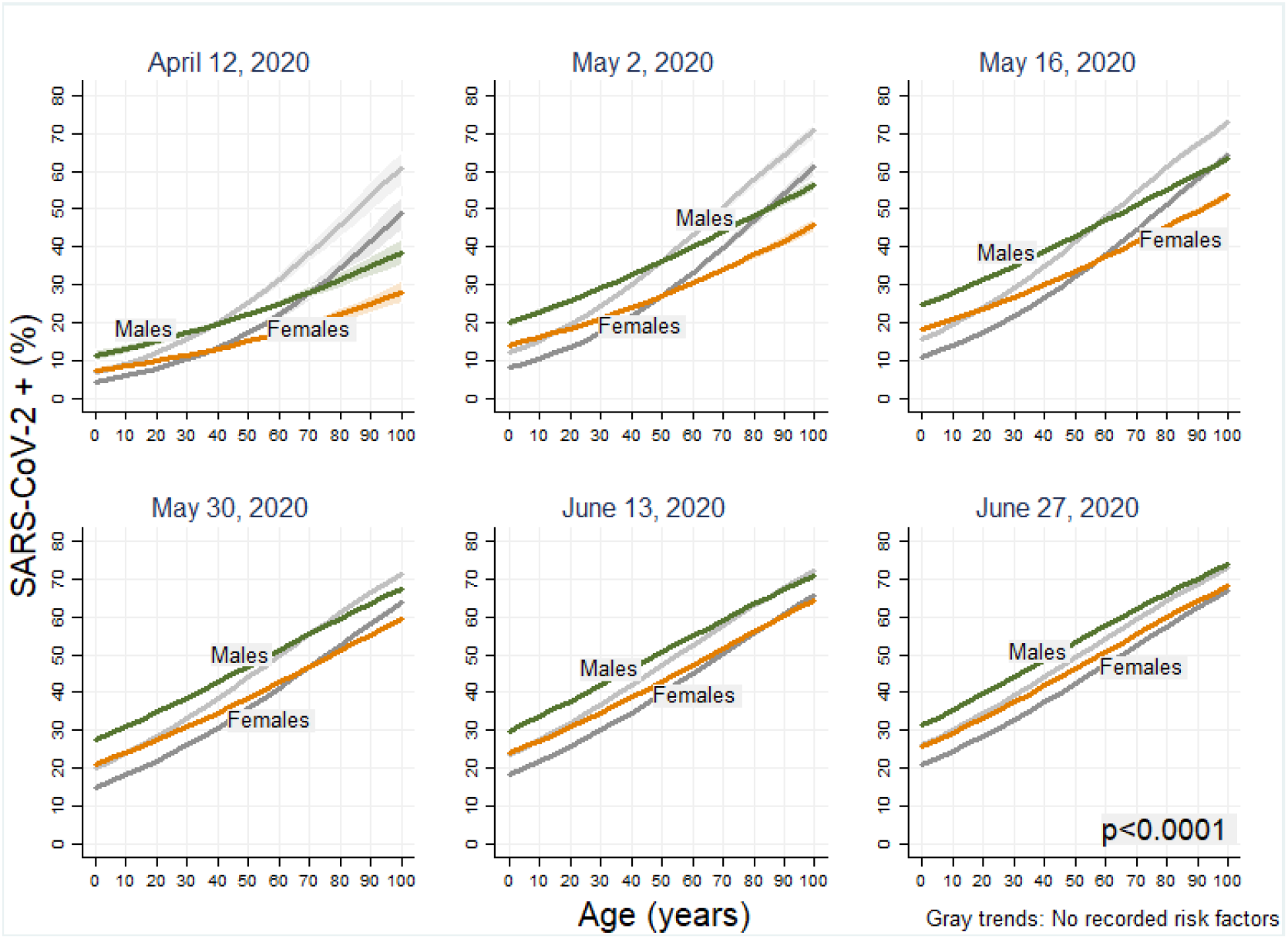
SARS-CoV-2 infection probability progression by date. Green and orange trends: Patients with at least one risk factor. Gray trends: Patients with no risk factors.

Our findings are in accordance to previous theories on the distribution of ACE2 receptors between age groups **(8)**, which lessen the probability of infection and later development of COVID-19 in younger people, and woman, who might benefit from hormonal downregulation of ACE-2 expression **(9)**. Structural factors might also decrease the chance of developing lung compromise as children’s upper airways increased resistance might lead to aerosol particles to deposit in the bronchi rather than alveoli **(10). Al**so, because children develop fewer complications and have a lower mortality than older patients, they are more likely to become intermediaries for viral spread even without manifest signs and symptoms **(11)**. Some limitations of this study include that we only examined data from Mexico because it is currently the only country that has an open data policy that includes a wide range of variables. Nevertheless, these results might reflect the situation of other Latin American countries with similar age, sex and comorbidity distributions.

In conclusion, we found our model could monitor accurately the probability of SARS-CoV-2 infection in relation to age, sex and the presence of at least one risk factor. Also, because the model can be applied to any particular political region within Mexico, it could help evaluate the contagion spread in specific vulnerable populations. Further studies are needed to determine the underlying nature of the mechanisms behind such observations.

## Data Availability

All data is available at https://www.gob.mx/salud/documentos/datos-abiertos-152127

## Declaration of conflicts of interest

We declare no financial, academic or institutional conflicts of interest that affected the conception, elaboration, execution and processing of this work.

ORCID:

- Juan Alonso Leon-Abarca:
- Esdras Ismael Musayón:

## Notes

### Competing Interest Statement

The authors have declared no competing interest.

### Funding Statement

No funding sources whatsoever

### Author Declarations

Declaration of conflicts of interest: We declare no financial, academic or institutional conflicts of interest that affected the conception, elaboration, execution and processing of this work.

